# Xuefu Zhuyu Oral Liquid as Adjunctive Therapy on Stable Angina: A Randomized Clinical Trial

**DOI:** 10.1101/2025.02.03.25321625

**Authors:** Wencong Cao, Shaojun Liao, Li Zhou, Geng Li, Junwen Jiang, Weihui Lv, Bo Dong, Li Liu, Jie Zhang, Yanchun Wang, Wenwei Ouyang, Yi Du, Zehuai Wen

## Abstract

**Background:** Stable angina (SA) is a leading cause of disability worldwide, and there is increasing interest in nonpharmacological treatments. Xuefu Zhuyu oral liquid (XZOL) is a Chinese medicine that has been approved in China for the treatment of SA, but further studies are needed to establish its efficacy and safety. We aimed to provide a reliable assessment of the safety and efficacy of XZOL in patients with SA.

**Methods:** We did a multi-centre, randomised, double-blind, placebo-controlled trial at 6 hospitals in China. Participants were randomized into 2 treatment groups, and allocated to receive of either XZOL or matching placebo over 12 weeks and were followed-up for 12 weeks. The primary efficacy outcome was the average pain intensity of angina, which was measured by the change from baseline to week 12 on a 10-cm visual analogue scale.

**Results:** Of 263 patients screened, 148 participants were randomly assigned with primary outcome data in the ITT (intention-to-treat) population (74 in the XZOL group and 74 in the placebo group), and 99 participants in the PP (per-protocol) population (48 in the XZOL group and 51 in the placebo group). Mean change in VAS at week 12 were −2.27 in the XZOL group and −1.85 in the placebo group (difference - 0.52, 95% CI −0.99 to −0.05; *P* = 0.029) among the ITT population, and −2.54 in the XZOL group and −1.79 in the placebo group (difference −0.81, 95% CI −1.40 to −0.22; *P* = 0.007) among the PP population. The neutral results were shown in adjusted and sensitivity analyses. There was no significant difference in adverse events.

**Conclusion:** This randomised, placebo-controlled, double-blind, clinical trial showed XZOL was associated with a significantly greater reduction in the levels of pain intensity of angina over 12 weeks and was superior to placebo in alleviating SA.

## 1. Introduction

Stable angina (SA), as a symptomatic problem precipitated by myocardial ischemia, is associated with an average annual risk of 3% to 4% for myocardial infarction or death (Joshi and de Lemos, 2021). A recent survey has shown a prevalence of SA of 9.6% in China (Greaves et al., 2014). Consequently, new demands arise for SA prevention, treatment, and the allocation of medical resources, considering China’s large population. Most interventions for SA target stopping or minimizing symptoms and improving quality of life and long-term morbidity and mortality. The options include lifestyle advice, drug treatment and revascularization (Henderson et al., 2012). However, due to the lack of obvious improvement to SA and side effects with limited antianginal drugs (Kones, 2010; Thadani, 2004) and percutaneous coronary intervention (Greaves et al., 2014), the majority of patients were still symptomatic and many of them perceived that SA had a major impact on their working life after receiving treatment (Manolis et al., 2019).

Chinese medicine has long been used in contemporary health care in China among patients with SA (Wang and Chen, 2018). There is now a considerable body of animal and early phase clinical data to support the use of various Chinese herbs to promote the prognosis and reduces the risk of cardiovascular events, benefiting patients with SA (Cai et al., 2018; Chen et al., 2021; De Biase et al., 2021; Lue et al., 2021; Wang et al., 2021; Wang et al., 2023; Yu et al., 2023). One classical Chinese herbal formula, Xuefu Zhuyu decoction, as documented in in “Yilin Gaicuo” by Wang Qingren during the Qing Dynasty. is a traditional herbal formula widely used in traditional Chinese medicine (TCM) for conditions associated with Qi stagnation and blood stasis, including chest pain, headache, palpitations, and insomnia, which are linked to cardiovascular and cerebrovascular disorders in modern medicine. Several experiments have provided evidence that Xuefu Zhuyu decoction protected cardiomyocytes against hypoxia/ reoxygenation injury by inhibiting autophagy (Shi et al., 2017) and demonstrated a superior ability to reverse myocardial fibrosis in spontaneously hypertensive rats (Zhang et al., 2016). What’s more, several small-scale trials have shown that Xuefu Zhuyu decoction is effective and safe for angina pectoris with ameliorating anginal symptoms, myocardial ischemia and fewer adverse effects (Liao et al., 2006; Liu et al., 2007; Liu et al., 2006; Zhang et al., 2021).

Xuefu Zhuyu oral liquid (XZOL), an improved dosage form of the decoction, is a patented drug made of eleven Chinese herbs, including Taoren (Semen Persicae), Honghua (Flos Carthami), Dihuang (Radix Rehmanniae), Danggui (Radix Angelicae Sinensis), Chuanxiong (Rhizoma Chuanxiong), Niuxi (Radix Achyranthis Bidentatae), Chishao (Radix Paeoniae Rubra), Chaihu (Radix Bupleuri Chinensis), Jiegeng (Radix Platycodi), Zhiqiao (Fructus Aurantii Submaturus) fried with bran, and Gancao (Radix Glycyrrhizae) (Supplementary file 1), and was approved by the China Food and Drug Administration in 2002 (Ref No. Z10950063). However, there has been no high-quality evidence of the efficacy and safety of XZOL for patients with SA. We therefore designed a 24-week randomized controlled trial to establish the efficacy and safety of XZOL in patients with SA.

## 2. Methods

### 2.1. Study Design

This multicenter, double-blind, randomized controlled trial was done between June 2020 and June 2022 at 6 sites in China. The aim was to assess the safety and efficacy of XZOL in patients with SA. The protocol was approved by the Ethics Committees at Guangdong Provincial Hospital of Chinese Medicine (No. BF2019-175-01) and registered with an identifier (ChiCTR1900026899) in the Chinese Clinical Trial Registry. We followed the Consolidated Standards of Reporting Trials Extension for Chinese Herbal Medicine Formulas 2017 reporting guideline (Cheng et al., 2017) for reporting this study. All participants provided written informed consent before participation. Additional details are presented in the study protocol (Liao et al., 2020).

### 2.2. Participants

Participants were adults aged 30 to 75 years. Eligible participants were the following: (1) who were diagnosed SA due to coronary heart disease by the Guideline for the Diagnosis and Treatment of Patients with Stable Ischemic Heart Disease issued by the Chinese Society of Cardiology in 2018 (Chinese Society of Cardiology, 2018) and the Guideline Update for the Management of Patients with Chronic Stable Angina Pectoris released by the American Heart Association/ American College of Cardiology in 2002 (Gibbons et al., 2003), (2) whose patterns were classified as Qi-stagnation and Blood-stasis pattern based on a validated diagnostic scale of Chinese medicine (Wang et al., 2018), (3) whose grading of angina of effort by the Canadian Cardiovascular Society (CCS) (Campeau, 1976) were either I, II, or III, (4) whose pain score on the visual analogue scale (VAS) ≥ 3 cm over the past 2 weeks. Key exclusion criteria included suffering from severe heart disease as well as any other disease that triggers chest pain, poorly controlled hypertension, or severe anxiety and depression (Zung Self-rating Anxiety Scale > 59 (Duan and Sheng, 2012; Zung, 1971) or Zung Self-rating Depression Scale > 62 (Duan and Sheng, 2012; ZUNG, 1965)). Details of the inclusion and exclusion criteria are provided in the appendix (Table 1, 2, and 3 in Supplementary file 2).

**Table 1.** Participant baseline characteristics (FAS)

| Variable | Participants, No. (%) (N = 148) |  |
| --- | --- | --- |
|  | XZOL group<br>(n = 74) | Placebo group<br>(n = 74) |
| Age, mean (SD), y | 61 (8.97) | 61 (8.61) |
| Gender, No. (%) |  |  |
| Male | 53 (71.6) | 48 (64.9) |
| Female | 21 (28.4) | 26 (35.1) |
| Height, mean (SD), cm | 167 (7.03) | 167 (6.93) |
| Weight, mean (SD), kg | 71.4 (11.41) | 70.2 (11.33) |
| BMI, median (SD), (kg/m<sup>2</sup>) | 25.4 (3.6) | 25.0 (3.1) |
| Race and ethnicity, No. (%) |  |  |
| Han | 72 (97.3) | 72 (97.3) |
| Not Han | 2 (2.7) | 2 (2.7) |
| Education, No. (%) |  |  |
| Less than high school up to completed high school | 62 (84.7) | 62 (84.7) |
| Incomplete college education or higher | 12 (16.2) | 12 (16.2) |
| Family history, No. (%) |  |  |
| Yes | 66 (89.2) | 63 (85.1) |
| No | 8 (10.8) | 11 (14.9) |
| Resting blood pressure, mean (SD), mm Hg |  |  |
| Systolic | 130 (9.74) | 129 (9.82) |
| Diastolic | 77 (8.08) | 78 (9.38) |
| Participant smokes any cigarette, No. (%) |  |  |
| Yes | 39 (52.7) | 27 (36.5) |
| No | 35 (47.3) | 47 (63.5) |
| Participant consumes alcoholic beverages, No. (%) |  |  |
| Yes | 24 (32.4) | 16 (21.6) |
| No | 50 (67.6) | 58 (78.4) |
| Disease duration, mean (SD), m | 44 (42.29) | 41 (51.04) |
| Previous use of PCI |  |  |
| Yes | 52 (70.3) | 54 (73.0) |
| No | 22 (29.7) | 20 (27.0) |

BMI = Body Mass Index; FAS = full analysis set; SD = standard deviation; PCI = percutaneous coronary intervention; XZOL = Xuefu Zhuyu oral liquid.

### 2.3. Randomization and masking

A center-stratified randomization sequence with a block of four was generated by SAS 9.2 (SAS Institute Inc., Cary, USA) and performed by the Institute of Basic Research in Clinical Medicine (IBRCM), China Academy of Chinese Medical Sciences (Beijing, China). After confirmation of eligibility, patients were randomly assigned (1:1) to either XZOL or matching placebo via the Interactive Web Response System developed by IBRCM.

The XZOL and the placebo were manufactured as oral liquids by Jilin Aodong Yanbian Pharmaceutical Co. Ltd. (Jilin, China) according to the requirements of good manufacturing practice. The placebo, instead of consisting of honey, white granulated sugar, fried brown sugar, ginseng essence, bitters, natural edible pigments, and food antiseptic, had good consistency with XZOL in appearance, color, taste, dosage, packaging, and labeling. Details are outlined in the supplementary file 1.

Participant allocation was concealed among during the trial period. The participants, investigators and statistician were unaware of the allocation and grouping, which IBRCM independently completed and managed.

### 2.4. Procedures

After enrolment, both participants in the XZOL group and the placebo group will be given conventional treatment for SA, following the Chinese Guidelines for Diagnosis and Treatment of Patients with Stable Coronary Artery Disease 2018 (Chinese Society of Cardiology, 2018). Nitroglycerin 0.5 mg can be taken sublingually when angina pectoris is intolerable. Participants in two groups received either the XZOL or the placebo, respectively, 20 ml/ time, 3 times per day, for 12 weeks. Follow-up evaluations were undertaken at the 2^nd^, 4^th^, 8^th^, 12^th^, and 24^th^ weeks after intervention (with a window of ± 2 to 7 days) by trained certified medical staff.

The trial was overseen by a steering committee and all serious adverse event reports were submitted to the ethics committee and the Data and Safety Monitoring Committee for review and monitoring. Demographic characteristics, including gender, race and ethnicity, were collected via the paper case report forms in reserved rooms, and then entered to the electronic data capture system developed by IBRCM. The quality of the data collection was monitored by the independent department of Jilin Aodong Yanbian Pharmaceutical Group Co. Ltd (Jilin, China).

### 2.5. Outcomes measurements

The primary outcome was the difference in average pain intensity of angina compared with baseline after 12 weeks of intervention, which was assessed using a 10-cm VAS. This VAS score was determined by measuring millimeters to the point marked by the participant (score range, 0 [no pain] to 10 cm [strongest pain], with lower scores indicating a better outcome according to participants’ perspective). Secondary outcomes were also assessed from baseline to the end of treatment (week 12) and the follow-up (week 24), including (1) angina attack frequency, (2) angina attack duration, (3) nitroglycerin dosage consumed, (4) angina severity by the CCS grading of effort angina, (5) Qi stagnation and Blood stasis pattern changes by the scale, (6) the impact of angina on the health status by the Seattle Angina Questionnaire (Liu, 2003), (7) quality of life by the EuroQol-5-Dimensions-5-Level (EQ-5D-5L) (Janssen et al., 2013), and (8) sleep quality by the Pittsburgh Sleep Quality Index (Buysse et al., 1989). All serious adverse events and adverse events (AEs) were recorded through to study completion on the case report forms, and regularly evaluated by the investigators. The causality between intervention and AEs was judged based on the WHO-UMC system for standardized case causality assessment (WHO, 2013).

### 2.6. Statistical analysis

According to the previous study (Yang et al., 2015), a sample of 61 participants per group was initially estimated to detect a superiority margin of 1.7 measured by VAS pain score, assuming a standard deviation of 1.5 with the test as 2-tailed, 90% power and a significance level of *P* < 0.05. An additional 20% drop-out was considered, totaling 152 participants, with 76 in each group.

The principal analysis used the full analysis set (FAS) from all randomly assigned patients who received at least one session of intervention and at least efficacy measure once (the intention-to-treat population). Missing data were filled by using multiple imputations. Continuous variables were reported as mean and standard deviation (SD) and compared with *t*-test or Wilcoxon rank-sum test as appropriate. Categorical variables were described as frequencies and percentages and compared with a Chi-squared test, Fisher’s exact test or Mann-Whitney *U* test as appropriate.

For the primary outcome, the differences from baseline in the VAS score at week 12 were analyzed by fitting a generalized linear model adjusting for the baseline VAS score, center, and interaction between center and treatment. For secondary outcomes, we verified statistically significant differences from baseline to week 24. AEs were analyzed using the safety set, comprising all post-randomization patients who received at least 1 session of intervention to assess the safety of the study drug.

We performed sensitivity analyses by replicating the analyses in the per-protocol set (PPS), which included all randomized patients who completed all of the intervention and assessments without serious protocol violations (per-protocol population). In addition, we performed the prespecified subgroup analyses by gender, age (> 65 years or ≤ 65 years), and CCS (Ⅰ and Ⅱ or Ⅲ) during the trial. These tests were followed by post hoc tests to identify significant differences at specific time points.

All analyses were conducted with a significance level of *P* < 0.05. Statistical analyses were performed using SPSS 18.0 (IBM SPSS Inc., Armonk, New York, USA) or SAS 9.2 (SAS Institute Inc., Cary, USA).

## 3. Results

From June 2020, to June 2022, of 263 patients screened, 152 were randomly assigned at 6 tertiary level hospitals in China. After excluding 4 people for whom no data of outcome assessment were available, 148 participants comprised the FAS with 74 participants randomized to the XZOL or the placebo group respectively. Among all patients, 99 completed all 12 weeks of intervention plus an additional 12 weeks of follow-up, including 48 patients in the XZOL group and 51 participants in the placebo group (Figure 1). Baseline characteristics of patients are shown in Table 1.

**Figure 1.**
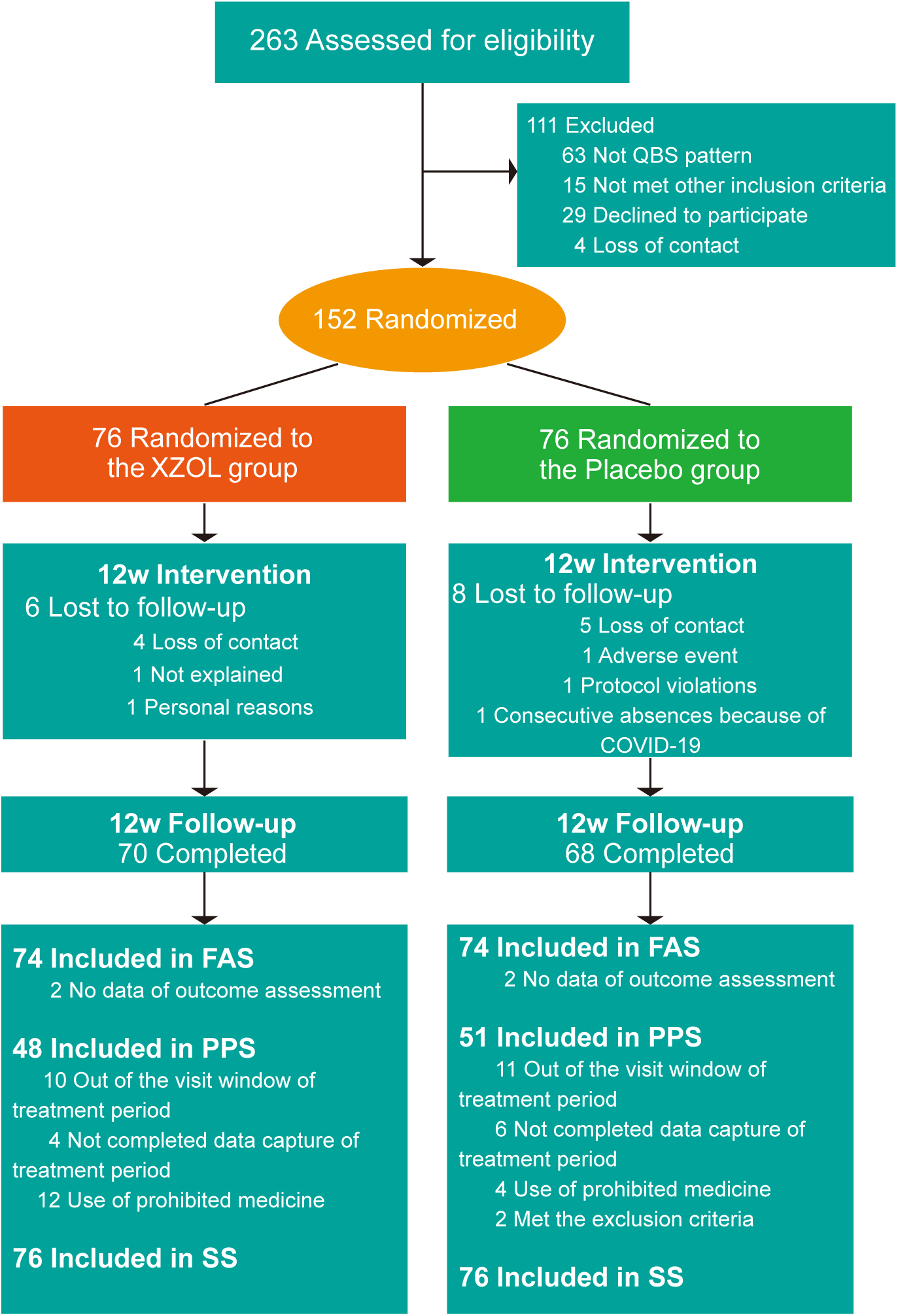
Flowchart of screening, randomization, and follow-up COVID-19 = Corona Virus Disease 2019; FAS = full analysis set; PPS = per-protocol set; QBS = Qi stagnation and Blood stasis; SS = safety set.

### 3.1. Primary outcome

Twelve weeks after randomization, lower VAS scores were observed in the XZOL group than that in the placebo group. A statistically significant difference (mean difference, −0.52 [95% CI, −0.99 to −0.05]; *P* = 0.029) in favor of the XZOL group was observed in the mean change in VAS from baseline to the end of treatment (week 12) (Table 2), even though it didn’t reach to the superiority margin of 1.7 as estimated.

**Table 2.**
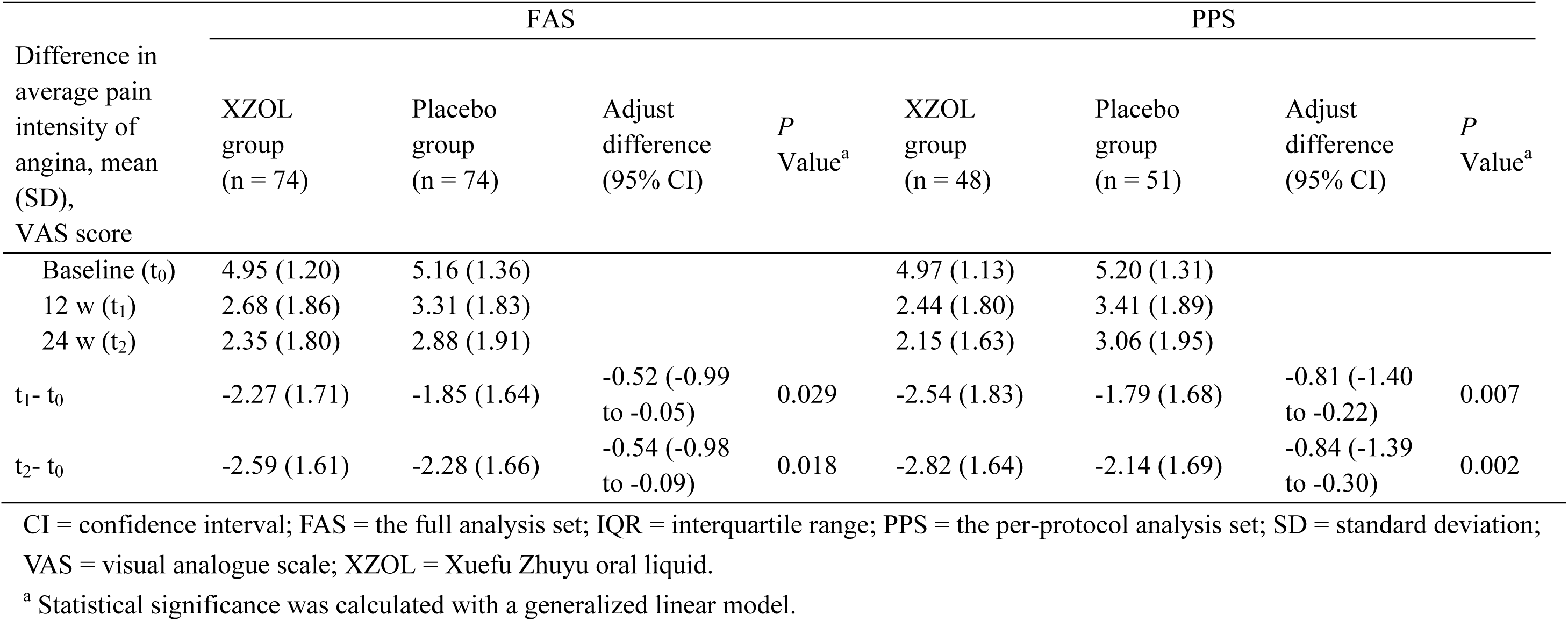
Primary outcomes.

For the sensitivity analyses, we found that the intergroup comparison results for the primary outcomes of the FAS were consistent with the results of the PPS (−0.81 [CI, −1.40 to −0.22]; *P* = 0.007) (Table 2).

In addition, changes in the mean VAS scores within the follow-up period differed significantly between the 2 groups at 24 weeks (Table 2). The trend of mean in VAS over time was similar between the 2 groups (Figure 2). The results of the PPS were consistent with those of the FAS.

**Figure 2.**
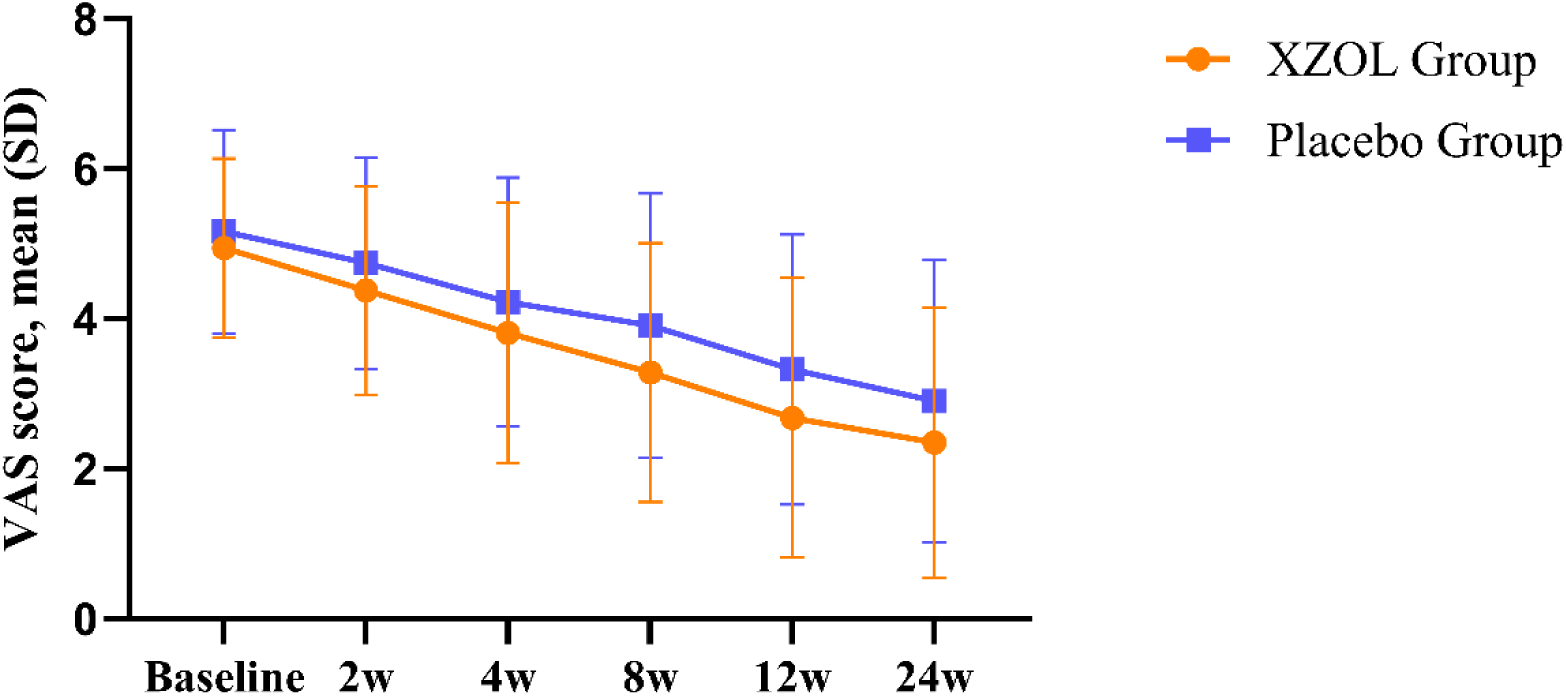
Difference in average pain intensity of angina over time during the study by FAS SD = standard deviation; VAS = visual analogue scale; XZOL = Xuefu Zhuyu oral liquid.

No significant differences were noted in the prespecified subgroup analyses by gender and age but by Ⅲ of CCS grading. A statistically significant difference in favor of the XZOL group was observed in CCS grading remission after 4 weeks (5.20 [1.47] in the XZOL group vs 6.68 [0.83] in the placebo group, −1.48 [CI, −3.31 to 0.35]; *P* = 0.040) (Online Table 4, 5, and 6 in Supplementary file 2).

### 3.2. Secondary outcomes

Regarding the FAS, a reduction was observed based on angina attack frequency and duration in both groups at each interview during the treatment and follow-up periods, but without statistically significant differences between them. Meanwhile, there was no difference in the number of patients using nitroglycerin in 2 groups during the 24-week study period, but not at twelve weeks (*P* = 0.013). CCS grading of effort angina was assessed at baseline, the end of treatment and follow-up periods. There were no significant differences in both groups in the proportion of patients with no change or with deterioration of Ⅰ (class I indicates angina with strenuous exertion) or more classes. Likewise, the proportions of patients with recovery and remission based on the Qi stagnation and Blood stasis pattern changes, Seattle Angina Questionnaire, EQ-5D-5L index, EQ-5D-5L VAS and the Pittsburgh sleep quality index were showed in both groups at each interview during the treatment and follow-up periods. However, there were no statistically significant differences between them. The per-protocol analysis showed the similar results as the FAS analysis (Table 3).

**Table 3.**
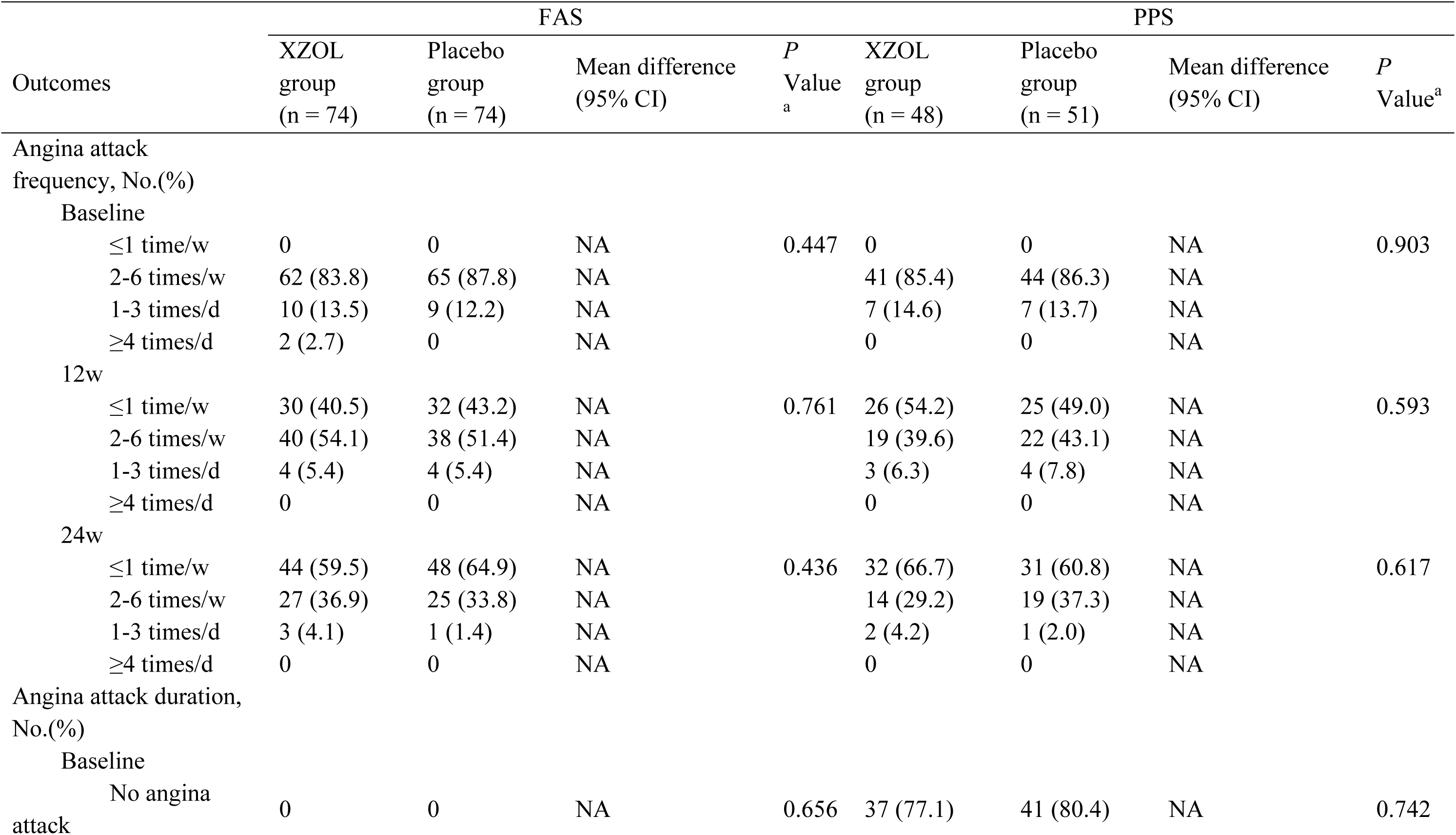
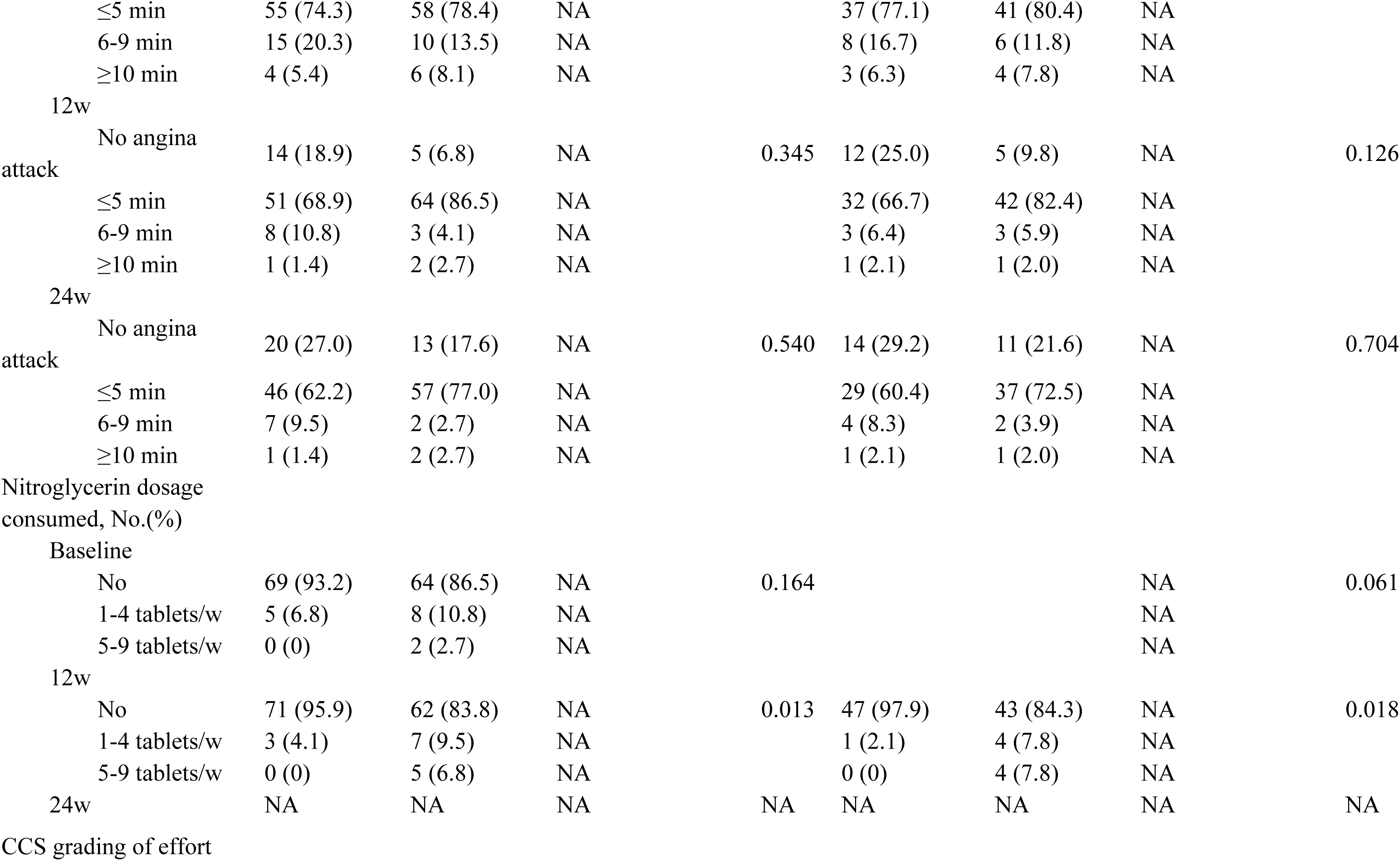
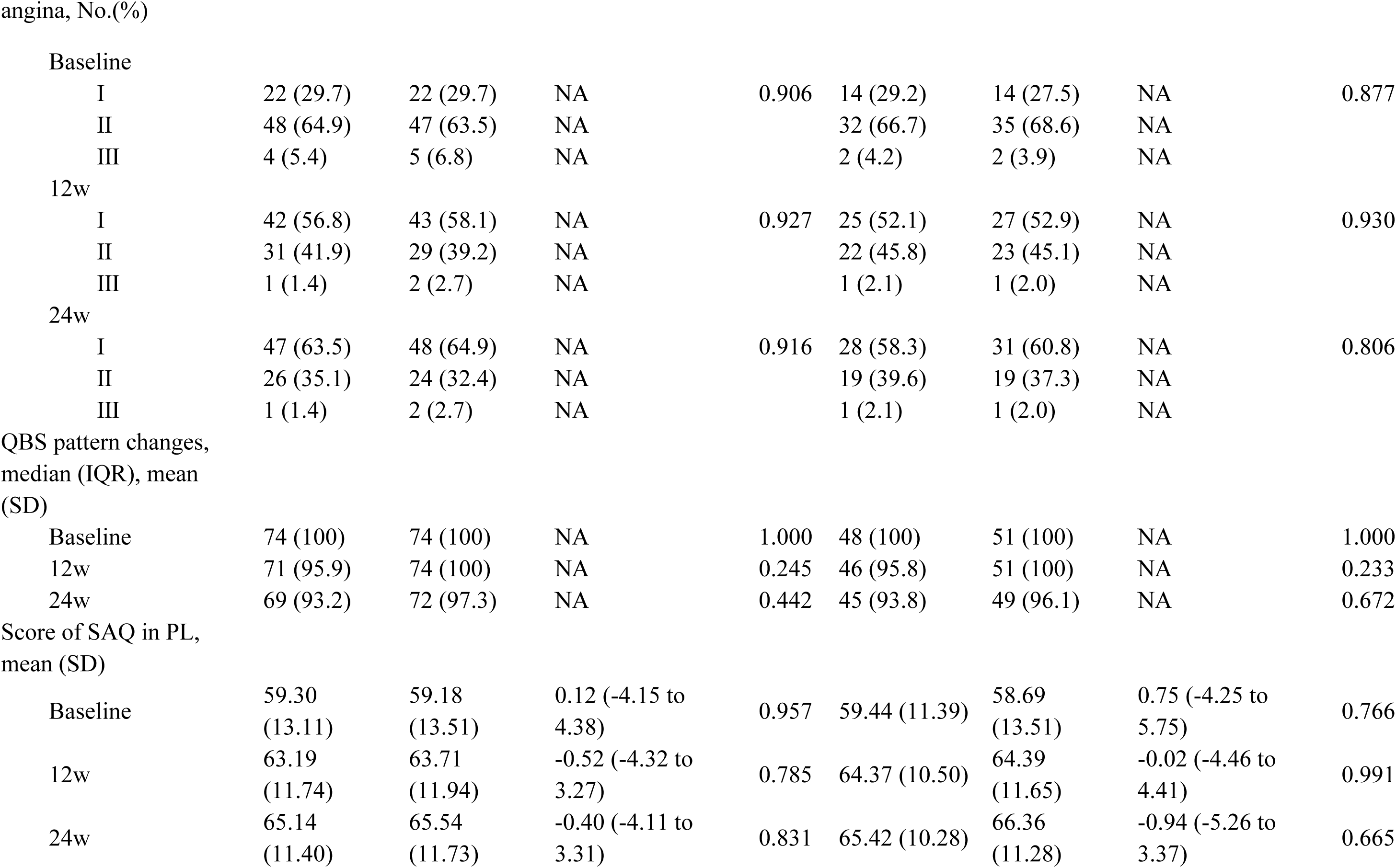
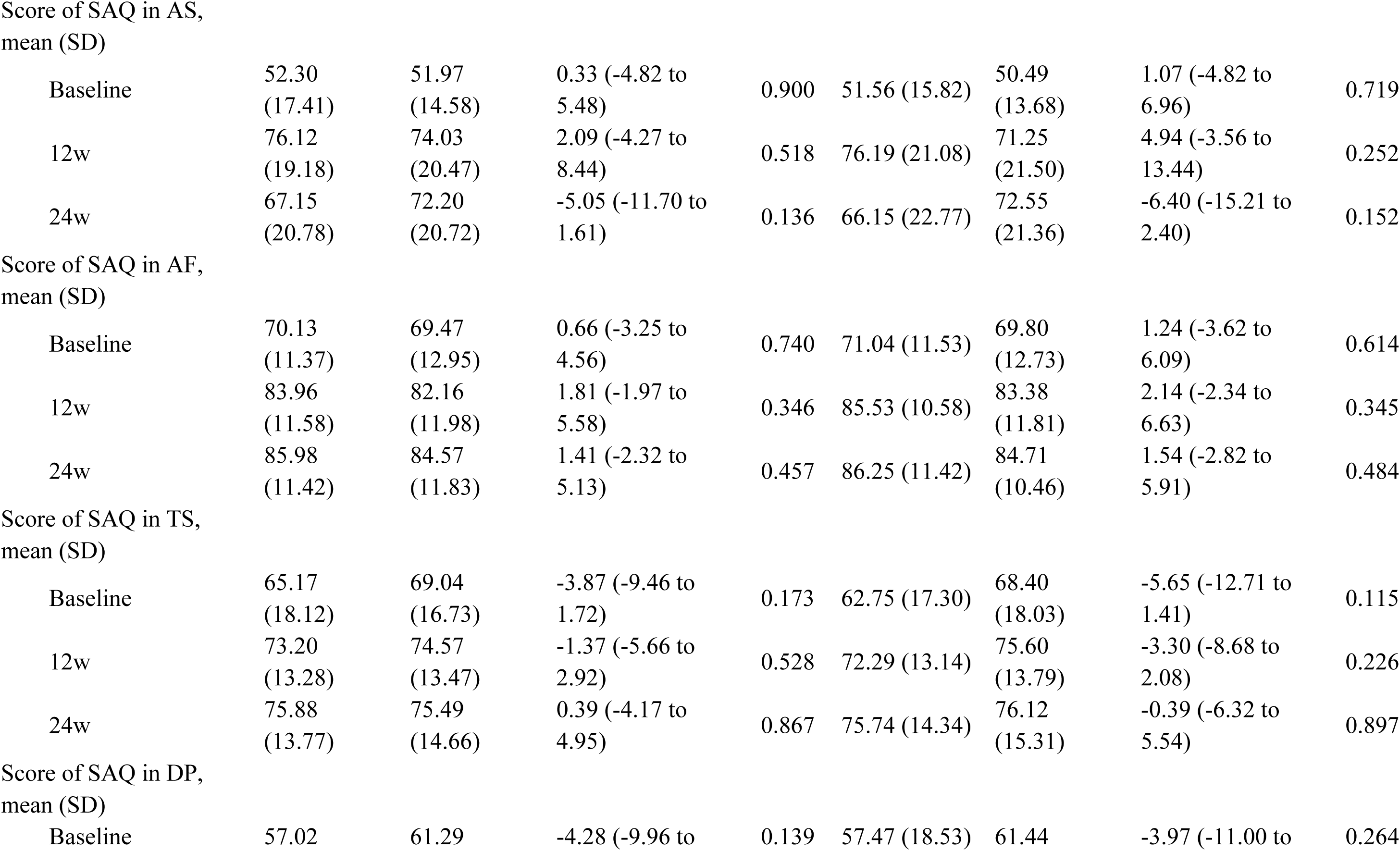
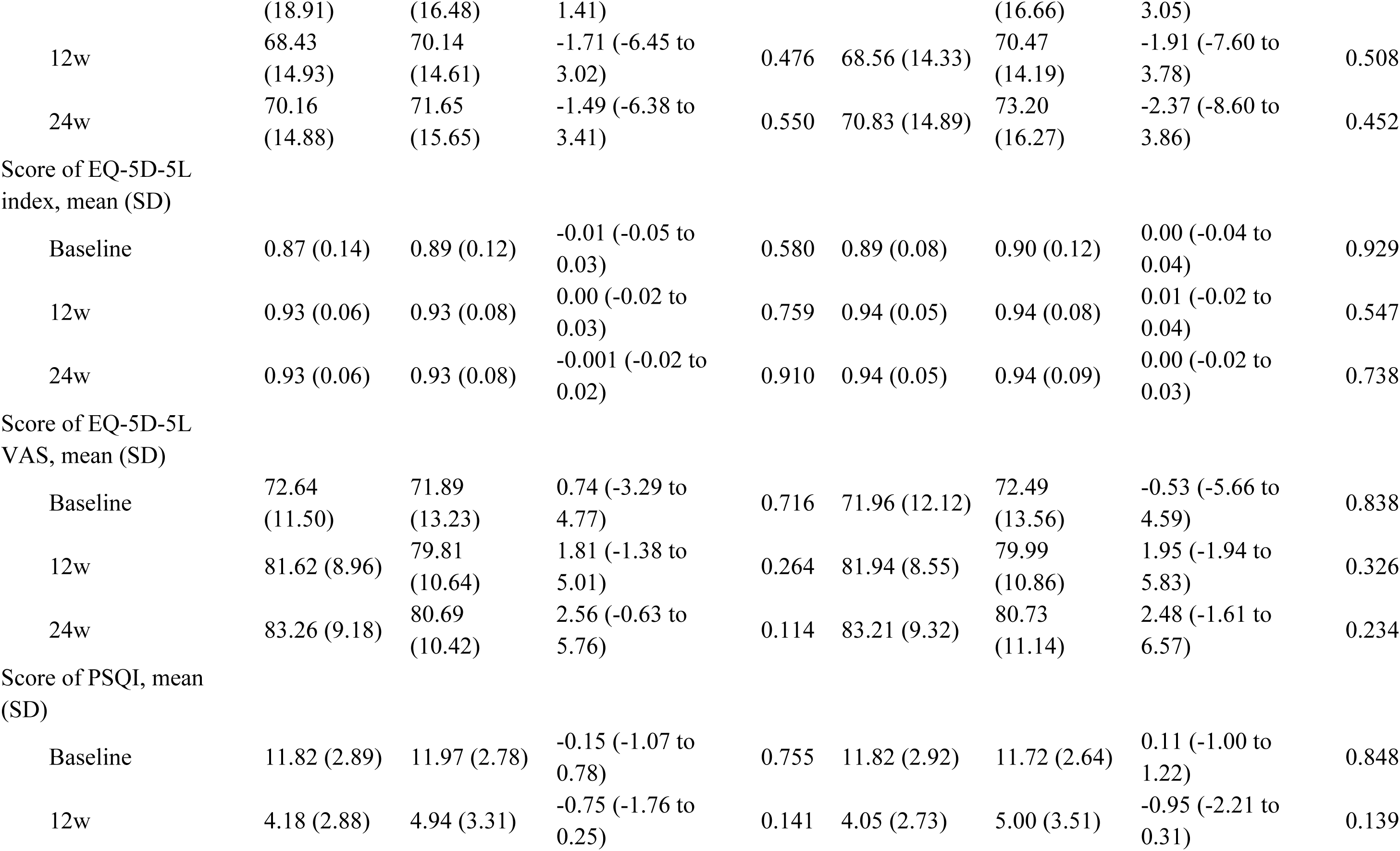
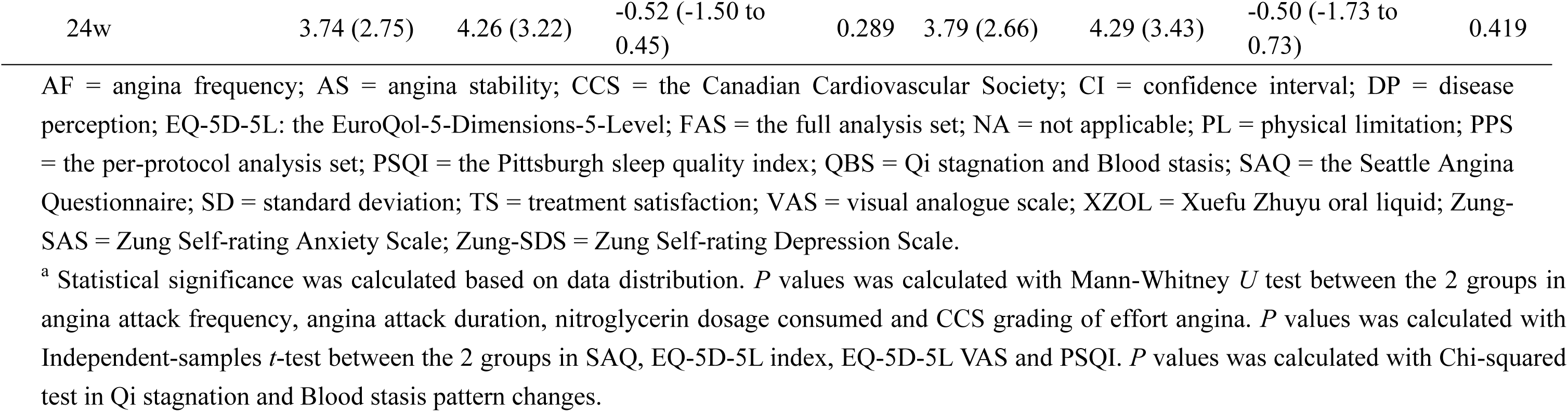
Secondary outcomes.

### 3.3 Adverse events

All AEs and adverse reaction (ADR) were observed and recorded in the trial. A total of 16 AEs occurred in 152 patients, including 8 (10.5%) in the XZOL group, as well as in the placebo group (Table 4), and no statistically significant difference was found between the groups in the proportion of participants with AEs (*P* = 1.000). A severe adverse event was recorded because 1 (1.3%) participant had unstable angina pectoris in the placebo group during the trial. But this was not considered related to the intervention, and this participant recovered fully from the AEs and remained in the trial. Only 1 participant in the placebo group dropped out due to stomachache (also reported as ADR), which was considered related to the intervention. And there was another ADR that 1 patient reported abnormal liver function in the XZOL group. This might be considered related to the intervention, and he recovered fully and remained in the trial. Other AEs were listed in Table 4.

**Table 4.**
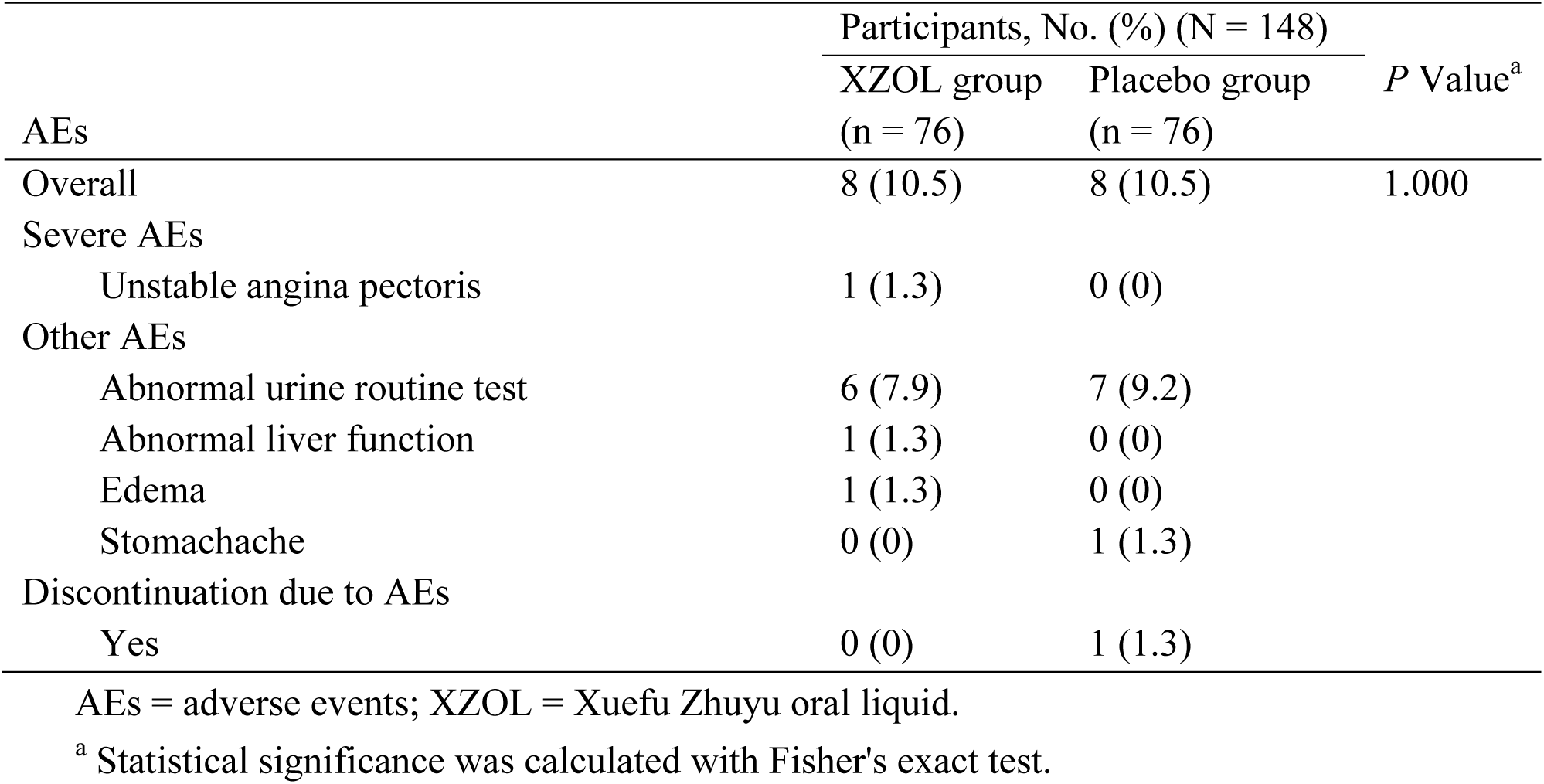
Adverse events listed by 2 groups.

| AEs | Participants, No. (%) (N = 148) |  | *P* Value<sup>a</sup> |
| --- | --- | --- | --- |
|  | XZOL group<br>(n = 76) | Placebo group<br>(n = 76) |  |
| Overall | 8 (10.5) | 8 (10.5) | 1.000 |
| Severe AEs |  |  |  |
| Unstable angina pectoris | 1 (1.3) | 0 (0) |  |
| Other AEs |  |  |  |
| Abnormal urine routine test | 6 (7.9) | 7 (9.2) |  |
| Abnormal liver function | 1 (1.3) | 0 (0) |  |
| Edema | 1 (1.3) | 0 (0) |  |
| Stomachache | 0 (0) | 1 (1.3) |  |
| Discontinuation due to AEs |  |  |  |
| Yes | 0 (0) | 1 (1.3) |  |

## 4. Discussion

This multicenter, randomized, controlled trial showed XZOL clinically relevant benefits in reducing the VAS scores to a greater degree than that in the placebo group after 12-week treatment. In addition, compared with the placebo, XZOL resulted in better remission of the VAS scores within the 12 weeks at the end of the follow-up period, namely effects lasted to 12 weeks after treatment ended. In other words, XZOL had superior to relieve the anginal symptoms. Furthermore, no significant differences were observed in the rates of AEs, evidencing practice safety. To our knowledge, the trial is the largest multicenter clinical trial to show the beneficial effect of XZOL as adjunctive treatment for SA within 24 weeks.

These preliminary findings are promising and support the adjunctive use of XZOL in SA treatment. Our findings are consistent with those of a previous systematic review (Yi et al., 2014) that examined the effectiveness of combined XZOL decoction and antianginal treatment vs antianginal medications alone. Anginal symptoms remission is crucial for patients, providing better quality of life and long-term prognosis, not just symptom improvement (Joshi and de Lemos, 2021). It is essential to highlight that Chinese medicine believes SA belongs to the categories of “heartache” and “chest stuffiness”, characterized by stabbing pain, chest tightness, and palpitation (Wang et al., 2021; Wang et al., 2020). We should underscore that in XZOL decoction, part of the effect is attributed to improving angina pectoris attack, increasing the levels of serum superoxide dismutase and interleukin-10 while simultaneously reducing the levels of serum soluble intercellular adhesion molecule-1, vascular endothelin-1 and interleukin-6 (Lai et al., 2023). This effect may reduce the inflammatory response. An additional hypothesis is that XZOL decoction may reduce platelet activating factor which would lead to the development of coronary atherosclerosis (Wen, 2017; Shao, 2020). An experimental study (Tang et al., 2014) confirmed this hypothesis, showing that the effect of XZOL decoction is possibly associated with restoring blood flow by improving the abnormal hemorheology of rabbits with blood stasis. Additionally, some studies observed that XZOL decoction can improve myocardial fibrosis (Jiang et al., 2018; Tang et al., 2015) and oxidative stress injury (Dou et al., 2019; Tang et al., 2018), which can partially reflect its effects of removing blood stasis and promoting blood circulation. XZOL, as an improved dosage form of XZOL decoction, strictly follows the original prescription and dosage. Therefore, it can be inferred that XZOL oral liquid could alleviate QBP through multiple approaches.

This study has strengths. First, the research was a rigorously designed, multicenter, and large-scale clinical trial, providing the reliable evidence for the clinical use of XZOL combined antianginal treatment for SA. In addition, the study included a more extended study period, which would help to better observe the XZOL’s prolonged effect at the 12-week follow-up. Our continuous follow-up also ensured patient compliance during the Corona Virus Disease 2019 epidemic.

The present study also has limitations. Firstly, the bias may be also introduced into the results by using self-report instruments to measure the outcomes. Other major limitations include the limited sample size and the relatively high loss of participants during the follow-up period. Additionally, the XZOL group with positive results indicates the need to investigate better the mechanisms of XZOL and nonspecific outcomes associated with SA.

## 5. Conclusions

Our study has shown that in patients with SA, use of XZOL over 12 weeks resulted in meaningful decrease in VAS pain score when compared with the placebo. We believe these findings support the consideration of XZOL as an option for adjunctive treatment in alleviating SA.

## Data Availability

The datasets used and analysed in the current study are available from the corresponding author on reasonable request.

## Authors’ contributions

ZHW and SJL was involved in study design and final approval of the manuscript. JWJ, WHL, BD, LL, JZ, YCW and YD were involved in data acquisition and interpretation. WCW was involved in the manuscript and revision. LZ, GL and WWOY were involved in data analysis and interpretation. All authors revised the manuscript critically and approved the final version of the manuscript.

## Ethics approval and consent to participate

All participants provided written informed consent and the protocol was approved by the Ethics Committees at Guangdong Provincial Hospital of Chinese Medicine (No. BF2019-175-01) and registered with an identifier (ChiCTR1900026899) in the Chinese Clinical Trial Registry.

## Consent for publication

Not applicable.

## Acknowledgments

We thank each of the study sites and principal investigators participating in this clinical trial for their outstanding conduct of the studies. We are also very grateful to all the patients who participated in these studies.

## Abbreviations

SA: stable angina
XZOL: Xuefu Zhuyu oral liquid
AEs: adverse events
FAS: full analysis set
CCS: Canadian Cardiovascular Society
VAS: visual analogue scale
IBRCM: Institute of Basic Research in Clinical Medicine
EQ-5D-5L: EuroQol-5-Dimensions-5-Level
PPS: per-protocol set
ADR: adverse reaction

## References

Buysse, D.J., Reynolds, C.F., Monk, T.H., Berman, S.R., Kupfer, D.J., 1989. The Pittsburgh Sleep Quality Index: a new instrument for psychiatric practice and research. Psychiatry Res. 28, 193–213.

Cai, X.M., Du, J., Li, L., Zhang, P.J., Zhou, H.N., Tan, X.M., Li, Y.B., Yu, C.Q., 2018. Clinical metabolomics analysis of therapeutic mechanism of Tongmai Yangxin Pill on stable angina. J Chromatogr B Analyt Technol Biomed Life Sci. 1100–1101, 106-112.

Campeau, L., 1976. Letter: Grading of angina pectoris. Circulation. 54, 522–523.

Chinese Society of Cardiology., 2018. Guidelines for the diagnosis and treatment of stable coronary artery disease. Chinese Journal of Cardiology. 46, 680–694.

Chen, Y., Xiao, X., Xu, X.L., Zhang, Z.P., Deng, Y., 2021. Traditional Chinese medicine in the prevention and treatment of stable angina pectoris in patients with coronary heart disease based on the theory of “phlegm and blood stasis” under guidance of evidence-based medicine: a prospective cohort study. J Tradit Chin Med. 41, 150–156.

Cheng, C.W., Wu, T.X., Shang, H.C., Li, Y.P., Altman, D.G., Moher, D., Bian, Z.X., CONSORT-CHM Formulas 2017 Group. 2017. CONSORT extension for Chinese herbal medicine formulas 2017: recommendations, explanation, and elaboration. Ann Intern Med. 167, 112–121.

De Biase, D., Biondi-Zoccai, G., Versaci, F., Frati, G., 2021. Management of chronic stable angina: modern microbiomedical research provides insights into traditional Chinese medicine treatments. J Cardiovasc Pharmacol. 77, 421–423.

Dou, X.B., Tang, H.Q., Zhao, Y.F., Zhao, Q.H., Li, K.M., Wang, L.Y., 2019. Effect of Xuefu Zhuyu Decoction on blood lipids and myocardial enzymology in animal model of coronary heart disease. Guangdong Medical Journal. 40(06):767–771.

Duan, Q.Q., Sheng, L., 2012. Differential validity of SAS and SDS among psychiatric non-psychotic outpatients and their partners. Chinese Mental Health Journal. 26, 676–679.

Gibbons, R.J., Abrams, J., Chatterjee, K., Daley, J., Deedwania, P.C., Douglas, J.S., Ferguson, T.J., Fihn, S.D., Fraker, T.J., Gardin, J.M., O’Rourke, R.A., Pasternak, R.C., Williams, S.V., Gibbons, R.J., Alpert, J.S., Antman, E.M., Hiratzka, L.F., Fuster, V., Faxon, D.P., Gregoratos, G., Jacobs, A.K., Smith, S.J., 2003. ACC/AHA 2002 guideline update for the management of patients with chronic stable angina-summary article: a report of the American College of Cardiology/American Heart Association Task Force on Practice Guidelines (Committee on the Management of Patients With Chronic Stable Angina). Circulation. 107, 149–158.

Greaves, K., Chen, Y., Appadurai, V., Hu, Z., Schofield, P., Chen, R., 2014. The prevalence of doctor-diagnosed angina in 4314 older adults in China and comparison with the Rose angina questionnaire: the 4 province study. Int J Cardiol. 177, 627–628.

Henderson, R.A., O, Flynn, N., 2012. Management of stable angina: summary of NICE guidance. Heart. 98, 500.

Janssen, M.F., Pickard, A.S., Golicki, D., Gudex, C., Niewada, M., Scalone, L., Swinburn, P., Busschbach, J., 2013. Measurement properties of the EQ-5D-5L compared to the EQ-5D-3L across eight patient groups: a multi-country study. Qual Life Res. 22, 1717–1727.

Jiang, X.J., Li, T., Wang, D.S., 2018. Protective effect of Xuefu Zhuyu decoction on blood stasis syndrome of coronary heart disease and influence of plasma contact activation system. Research and Practice on Chinese Medicine. 32 (05): 19–22.

Joshi, P.H., de Lemos, J.A., 2021. Diagnosis and Management of Stable Angina: A Review. JAMA. 325, 1765–1778.

Kones, R., 2010. Recent advances in the management of chronic stable angina II. Anti-ischemic therapy, options for refractory angina, risk factor reduction, and revascularization. Vasc Health Risk Manag. 6, 749–774.

Lai, J., Wang, M., Deng, L., Wu, W.F., Meng, Q.W., 2023. Effects of Xuefu Zhuyu decoction on SOD, ET-1, sICAM-1 and Th1/Th2 balance in patients with stable angina pectoris of coronary heart disease differentiated as phlegm and blood stasis obstruction type. Journal of Guangzhou University of Traditional Chinese Medicine. 40(4):820–826.

Liao, S.J., Zhang, Z., Li, G., Zhou, L., Jiang, J.W., Zhang, N., Wang, Y., Du Y., Wen, Z.H., 2020. Chinese herbal formula Xuefu Zhuyu for stable angina (CheruSA): study protocol for a multicenter randomized controlled trial. Evid Based Complement Alternat Med. 7612721.

Liao, Z.H., Liang, Y., 2006. Effect of Xuefuzhuyu Oral Solution (XFOS) on coronary heart disease with angina pectoris. Medicine Industry Information. 3(05): 17–18.

Liu, J.G., Xu, H., Dong, G.J., Shi, D.Z., 2007. Effect of Xuefu Zhuyu Oral Liquids on platelet activating factor in patients with angina pectoris due to coronary heart disease. Journal of Changchun University of Traditional Chinese Medicine. 23(1): 29–31.

Liu, J.G., Xu, H., Dong, G.J., Shi, D.Z., Zhou, G.H., Zhu, J.H., Li, T., 2006. Xuefuzhuyu oral liquid on vascular endothelium function and hemorrheologic influence in patients with angina pectoris and blood-stasis syndrome. Chinese Journal of Integrative Medicine on Cardio-/Cerebrovascuiar Disease. 4(8): 659–661.

Liu, S.H., 2003. The evaluation of the reliability and validity of SAQ applied in a group of Chinese patients with CHD. Thesis of Master Degree. Tianjin Medical University. 53.

Lue, H.C., Su, Y.C., Lin, S.J.S, Huang, Y.C., Chang, Y.H., Lin, I.H., Yang, S.P., 2021. Taipei consensus on integrative traditional Chinese and Western Medicine. Journal of the Formosan Medical Association. 120(1): 34–47.

Manolis, A.J., Ambrosio, G., Collins, P., Dechend, R., Lopez-Sendon, J., Pegoraro, V., Camm, A.J., 2019. Impact of stable angina on health status and quality of life perception of currently treated patients. The BRIDGE 2 survey. Eur J Intern Med. 70, 60–67.

Shao, Z.M., 2020. The observation on clinical effect of modified xuefuzhuyu decoction on coronary heart disease induced angina pectoris and the impact on platelet activating factor (PAF). Clinical Research. 28(2):126–127.

Shi, X.W., Zhu, H.Y., Zhang, Y.Y., Zhou, M.M., Tang, D.L., Zhang, H.M., 2017. XuefuZhuyu decoction protected cardiomyocytes against hypoxia/reoxygenation injury by inhibiting autophagy. BMC Complement Altern Med. 17, 325.

Tang, H.Q., Pang L.L., Zhang, S.T., Feng, Y., Ning, W.L., Wang, L.Y., 2018. Effect of Xuefuzhuyu decoction on oxidative stress in rabbits with blood stasis syndrome of coronary heart disease. Progress in Veterinary Medicine. 39(8):31–35.

Tang, H.Q., Zhao, S.M., Huang, J.J., Mo, X.Q., Wang, L.Y., Wang, J.H., Lian, C.R., 2014. Effects of Xuefuzhuyu decoction formula on hemorheology in model rabbits with coronary heart disease at stage of blood stasis sybdrome. Progress in Veterinary Medicine. 35(3):44–47.

Tang, H.Q., Zhao, S.M., Huang, J.J, Wang, J.H., Wang, B., Mo, X.Q., Wang, L.Y., Lian, C.R., 2015. Effect of Xuefuzhuyu decoction on cardia function, cardiac muscle and blood vessel in rabbit with coronary heart disease at stage of blood stasis syndrome. Chinese Journal of Experimental Traditional Medical Formulae. 21(02):165–169.

Thadani, U., 2004. Current medical management of chronic stable angina. J Cardiovasc Pharmacol Ther. Sep:9 Suppl 1:S11–29; quiz S98-9.

Wang, D.Y., Li, C.Y., Xu, X.Q., Xu, H., Guo, C.C., Wang, J.P., Guo, J.Y., Huang, L., 2021. Effect of Yugengtongyu Granules in patients with stable coronary artery disease on reducing adverse cardiovascular events: a double-blind controlled trial. J Altern Complement Med. 27, 142–149.

Wang, J., Chen, G., 2018. The consensus of Chinese experts on the diagnosis and treatment of coronary heart disease with stable angina pectoris by traditional Chinese medicine. J Tradit Chin Med. 59(5): 447–450.

Wang, J., Gao, J.L., Chen, G., He, H.Q., 2018. Constructing diagnosis scale for qi stagnation and blood stasis syndrome. Chinese Journal of Experimental Traditional Medical Formulae. 24, 16–20.

Wang, W., Cao, L., Ren, P., Zhu, B.B., Liu, K., 2023. Network Meta-analysis of Chinese medicine injection combined with conventional western medicine in treatment of stable angina pectoris. Zhongguo Zhong Yao Za Zhi. 48, 1652–1663.

Wang, X.R., Song, D.D., Tao, T.Q., He, T., Wu, X.D., Li, X.M., Liu, X.H., 2021. Qi-regulating and blood circulation-promoting therapy improves health status of stable angina pectoris patients with depressive symptoms. Evid Based Complement Alternat Med. 7319417.

Wang, Y., Zhang, L., Pan, Y.J., Fu, W., Huang, S.W., Xu, B., Dou, L.P., Hou, Q., Li, C., Yu, L., Zhou, H.F., Yang, J.H., Wan, H.T., 2020. Investigation of invigorating Qi and activating blood circulation prescriptions in treating Qi deficiency and blood stasis syndrome of ischemic stroke patients: study protocol for a randomized controlled trial. Front Pharmacol. 11, 892.

Wen, H.J., 2017. Clinical effect of modified xuefuzhuyu decoction on coronary heart disease induced angina pectoris and the impact on platelet activating factor. Practical Journal of Cardiac Cerebral Pneumal and Vascular Disease. 25(12):83–86.

WHO. The use of the WHO-UMC system for standardised case causality assessment. 5 June 2013. WHO. Available at: https://www.who.int/docs/default-source/medicines/pharmacovigilance/whocausality-assessment.pdf. Accessed May 30, 2024.

Yang, S.Q., Lv, X.M., Sun, J.H., Liu, W.H., 2015. Effect of Xuefu Zhuyu Decoction of angina pectoris due to coronary heart disease. Pharmacology and Clinics of Chinese Materia Medica. 31, 144–146.

Yi, G.Z., Qiu, Y.Q., Xiao, Y., Yuan, L.X., 2014. The usefulness of xuefu zhuyu tang for patients with angina pectoris: a meta-analysis and systematic review. Evid Based Complement Alternat Med. 521602.

Yu, L.H., Wang, Z.H., Xu, C.X., Liu, A.X., Li, T., Wang, Y.B., Lu, X.Y., Xu, H., 2023. Integrated Chinese and Western medicine for stable angina pectoris of coronary heart disease: a real-world study including 690 patients. Front Cardiovasc Med. 10, 1194082.

Zhang, G.H., Yang, G., Deng, Y., Zhao, X.L., Yang, Y.B., Rao, J.J., Wang, W.Y., Liu, X., He, J., Lv, L., 2016. Ameliorative effects of Xue-Fu-Zhu-Yu decoction, Tian-Ma-Gou-Teng-Yin and Wen-Dan decoction on myocardial fibrosis in a hypertensive rat mode. BMC Complement Altern Med. 16, 56.

Zhang, S., Chen, Z.L., Tang, Y.P., Duan, J.L., Yao, K.W., 2021. Efficacy and Safety of Xue-Fu-Zhu-Yu Decoction for Patients with Coronary Heart Disease: A Systematic Review and Meta-Analysis. Evid Based Complement Alternat Med. 9931826.

Zung, W.W., 1965. A sself-rating depression scale. Arch Gen Psychiatry. 12, 63–70.

Zung, W.W., 1971. A rating instrument for anxiety disorders. Psychosomatics. 12, 371–379.

